# Loss to follow-up of ambulatory patients in the transition to telemedicine in the COVID-19 pandemic at a reference center for mental health in Lima, Peru

**DOI:** 10.1101/2024.08.27.24312689

**Authors:** Paulo Ruiz-Grosso, Abel Sagastegui, Sonia Zevallos-Bustamante

## Abstract

**Problem Statement:** During the COVID-19 pandemic, health care services were limited by the restrictive measures implemented. As an adaptation mechanism, telemedicine was introduced for ambulatory care at the Honorio Delgado Hideyo Noguchi National Institute of Mental Health (NIMH). This study aimed to estimate the survival function (SF) for loss to follow-up (LTFU) over two years before and after the onset of the COVID-19 pandemic, and its association with clinical and sociodemographic variables.

**Study Design:** A single-cohort study was conducted, following a random sample of adult ambulatory patients at NIHM from April 15, 2018, to April 15, 2022. Patients were followed until LTFU, referral to another institution, death, or the end of study. The main analysis involved estimating the SF for LTFU for the overall follow up period, as well as separately for the periods pre and post implementation of telemedicine. Also, risk factors hypotheses were tested using Cox’s regression.

**Results:** Data from 4887 visits of 356 patients were collected. A total of 118 (33.1%) presented LTFU, with SF of 53.9% during the overall four years of follow-up. After two years of follow-, those starting treatment at NIMH before the implementation of telemedicine had a higher SF (77.3 vs 63%). A higher hazard ratio (HR) for LTFU was found in the group that started care at NIMH after the implementation of telemedicine, compared to those who started previously (HR=2.53; 95% CI: 1.55-4.51). Additionally, receiving care in the anxiety disorders (HR=1.86; 95% CI: 1.03-3.33) and personality disorders programs (HR=1.81; 95% CI: 1.02-3.22) was associated with a higher risk of LTFU compared to the psychosis program. No significant difference was found in the risk of LTFU between telemedicine vs. face-to face attention.

**Conclusions:** A significant association was found between LTFU and starting treatment at NIMH after the onset of the COVID-19 pandemic and implementation of telemedicine. However, no evidence supports that this might be due to the practice of telemedicine. A different clinical profile of patients that started treatment at NIMH following the government implementation of changes to the public health system might explain these findings and should be studied.

**Significant Outcomes:**

- A moderate association was found between starting treatment at NIMH and loss to follow up in the period after the implementation of telemedicine due to the onset of COVID-19.
- This, however, does not seem to be related to the practice of telemedicine, but to changes in the characteristics of patients across pre and post COVID 19.

**Limitations:**

- The reason for loss to follow up (death, return to a lower-level health service, improvement of symptoms, etc.), our main outcome variable, could not be determined using the clinical and administrative data available.

## Introduction

The COVID-19 pandemic resulted in a significant challenge to public health systems worldwide, especially in developing countries [1–3]. The uncertainty, increased stress related to health issues that characterized this period saw an increase in the development of symptoms of depression, anxiety and sleep disorders in different populations such as adults, children and adolescents and elderly [4–10]. Also, there was concern for the continuation of care of patients on chronic antipsychotic and antidepressant treatments that were known to be at higher risk of severe illness from SARS-CoV-2, as they frequently present comorbid metabolic disorders and greater difficulties with self-care and proper application of hygiene measures [11,12].

The control measures adopted by governments around the world ranged from case tracing to prolonged quarantines, aiming to decrease the transmission of SARS-CoV-2. The Peruvian government adopted one of the longest and widespread mandatory social isolation, starting on March 15^th^, 2020; and reported one of the highest per capita mortality rates globally [1,13,14]. At the same time, most health facilities from first to third (highest) level of attention were mandated to suspend any service that was not considered an emergency. The National Institute of Mental Health “Honorio Delgado-Hideyo Noguchi” (NIMH) complied with this measure, starting the use of telemedicine for ambulatory patients.

Before the onset of the COVID-19 pandemic, telemedicine in psychiatry was studied, showing good consistency in the assessments between remote and face-to-face attention [15,16]. Some research showed that the adaptation of regulations and administrative mechanisms seemed to be the main barriers to adopting telemedicine in psychiatry. Even in developed countries, adopting this form of care appeared to improve service volume and reduce barriers to receiving care [17–19]. However, in countries with more variation in the development of health systems, the integration of this modality has been more variable [18].

These findings have been found consistent in studies performed during the COVID-19 pandemic [20,21]. In Peru, *Gayoso et al*. reported that most patients surveyed from a third level hospital where they received psychiatry consultation via telemedicine perceived improved health access (79.8%), time savings (89.6%), and found the procedure simple (80.3%) and comfortable to use (72.3%). The number of electronic devices and education level were positively associated with higher satisfaction, with a high proportion expressing a desire to continue this type of consultation [22]. From the caregiver’s point of view, *Valle et al*. reported results from a study that collected data immediately after the implementation of telemedicine in another specialized health center in Lima, Perú. This report showed that the majority (68.6%) identified teleconsultations as a quick form of care and found that time was generally not an issue (77.1%). On the other hand, most psychiatrists surveyed (57.1%) had no prior experience with telemedicine before the pandemic, there was no established protocol for this type of care in their institution (62.9%), and they found it problematic not to access the patient’s medical record (71.4%). Most proposed improvements on infrastructure and establishing a standardized workflow [23].

Despite the evidence in favor of the use of telemedicine, some important outcomes, such as loss to follow up (LTFU) was not reported for populations with a higher degree of severity of mental disorders, and, as far as we know, there is no study that compared this outcome in scenarios, pre and during the onset of the COVID-19 pandemic. In this study, we aim to estimate the survival function for LTFU in ambulatory patients during the period from April 15, 2018, to April 15, 2022; a period that includes two years before and after the start of telemedicine care at the NIMH. Additionally, we aim to identify sociodemographic and clinical variables that might modify the risk for LTFU during this period.

## Methods

### Study Design

This research followed a single cohort study design. Clinical data from adult ambulatory patients was collected from April 15^th^, 2018, two years before the start of use of telemedicine due to COVID or the date of first ambulatory consultation afterwards. Follow up finished in the date of the last consultation before patients were LTFU, were reported as deceased in any section of the clinical chart, or April 15^th^, 2022, when the study period ended. This design allowed to collect information in two different periods, the first, a pre-pandemic period (from April 15^th^, 2018, to April 15^th^, 2020) in which telemedicine was not used for ambulatory consultation; and a pandemic period (from April 15^th^, 2020, to April 15^th^, 2022), in which telemedicine was used as the only mode of attention for ambulatory patients, due to the nationwide adaptation strategies used to manage the COVID pandemic.

Data analysis followed a time to event approach, in which the main event of interest was LTFU. We estimated survival function for LTFU during the total follow-up period and for the pre-pandemic and pandemic periods. Afterwards Cox regression was used to assess the association of clinical variables, including type of attention (telemedicine vs. face to face), period in which each participant started attention at the NIMH-P, disease group, as well as other common sociodemographic variables such as age, sex, marital and education status.

### Sample Design

The population under study included adult ambulatory patients at the National Institute of Mental Health “Honorio Delgado – Hideyo Noguchi” (NIMH) that registered at least one appointment between April 15, 2018, and April 15, 2022. The NIMH is a specialized institution that acts as the reference center for the public health system in the northern region of Lima, capital of Peru; and provide ambulatory, hospitalization, rehabilitation and emergency services. During the follow up, we can identify two different periods. From April 15^th^, 2018, to April 15^th^, 2020, ambulatory attention was provided only in the traditional, face-to-face mode; and from April 15^th^, 2020, to April 15^th^, 2022, in which, as part of the country-wide adaptations to the COVID pandemic, telemedicine became the only mode for ambulatory attention for at least until the early days of 2022, in which face-to-face ambulatory care was progressively re-implemented.

In this study, we included adult (18-year-old or older) ambulatory patients that had at least one scheduled appointment with a psychiatrist during the follow up period, and their main diagnosis was of any psychosis, affective, anxiety and personality disorders. No exclusion criteria were applied. Eligible individuals were randomly selected from a list provided by the statistics department of NIMH until sample size was achieved. Data was collected from both physical and electronic clinical charts using Google Forms. Data form 375 individuals were required to detect a difference of at least 15% in the survival function between two categorical variables, assuming 5% types I and 20% type II error probability.

## Measures

### Outcome Variable

The main outcome variable was loss to follow-up (LTFU) and was defined as missing an appointment for a scheduled ambulatory attention during the follow-up period, and not attending any scheduled appointment thereafter. If the clinical chart showed information that the patient stopped attending their medical appointments due to medical indication (such as being discharged), informing of their deceased, or was transferred to another medical center to continue their treatment, this was not considered as LTFU.

### Sociodemographic and clinical covariates

Sociodemographic variables included age, gender (male/female), educational attainment (less than elementary/complete elementary/complete high school/complete and incomplete higher education), marital status (single/married/cohabitant/divorced/widow). Clinical covariates included the program of care (psychoses, elderly, affective, anxiety and personality disorders), mode of first admission to INSM (ambulatory/emergency), date of first attention at INSM (previous or after April 15^th^, 2020), period of attention (previous or after April 15^th^, 2020), mode of appointment (ambulatory face-to-face/ambulatory telemedicine/emergency), severity of evolution (no change, improvement, worsening).

## Data analysis

Data analysis followed the usual process of exploring and describing variables individually as proportions and means; then, we estimated the survival function for LTFU during the entire follow up period and then for those that started their attention at NIMH before and after April 15^th^, 2020 (start of the use of telemedicine). Bivariate and multivariate analyses were performed using Cox regression and stratification was further used to explore differences in magnitude of hazards by key clinical variables, including period of initial treatment at NIMH and program care.

## Ethical considerations

This study received approval from the NIMH Institutional Review Board before data collection started. To preserve the confidentiality of the information, a database linking the clinical chart numbers with an alphanumeric code was created, separated from the data to be analyzed. Only the main investigators had access to the database with the identities, and this information was used only to verify information during the data analysis.

## Results

### Sample characteristics

Data of 4,887 ambulatory attentions, corresponding to 356 individuals was collected between April 15, 2018, and April 15, 2022. The mean age in the sample was 36.7 years (SD=17.3), with 52.9% of the patients being female. The most common level of education was complete high school (41.8%), followed by complete elementary school (22.2%) and complete higher education (18.8%). Also, 77.7% were single, and 18.6% were married or cohabiting. The psychosis program provided care for 41.4% of patients, the affective disorders program for 20.8%, the anxiety disorders program for 11.8%, the personality disorders program for 12.9%, and the older adults program 12.9% of patients.

A total of 270 (76.1%) patients had their first attention at NIMH before April 15^th^, 2020; while 85 (23.9%) started their treatment at NIMH later. The type of first attention at NIMH was ambulatory for 149 (65.1%) patients, while 77 (33.6%) patients received the first attention at the emergency department. During follow-up, data were collected from a total of 1,933 (39.6%) face-to-face ambulatory consultations, 2,281 (46.7%) telemedicine ambulatory consultations, and 668 (13.7%) emergency consultations. Finally, 118 (33.1%) of 356 patients experienced LTFU during the study period. Details in Table 1.

**Table 1.** Sample characteristics.

| **Variable** | **n** | **%** |
| --- | --- | --- |
| <u>*Sex*</u> |  |  |
| Female | 188 | 52.9 |
| Male | 167 | 47.1 |
| Age* | 356 | 38.7 (17.3) |
| <u>*Instruction attained*</u> |  |  |
| Less than complete elementary school | 23 | 6.5 |
| Complete elementary school | 78 | 22.2 |
| Complete high school | 147 | 41.8 |
| Complete higher education | 66 | 18.7 |
| Non-complete higher education | 38 | 10.8 |
| <u>*Marriage status*</u> |  |  |
| Single | 275 | 77.7 |
| Married | 48 | 13.6 |
| Cohabitant | 18 | 5.1 |
| Divorced | 6 | 1.7 |
| Widowed | 7 | 1.9 |
| <u>*Program of care*</u> |  |  |
| Psychoses | 147 | 41.4 |
| Affective disorders | 74 | 20.8 |
| Anxiety disorders | 42 | 11.8 |
| Personality disorders | 46 | 13.0 |
| Elderly | 46 | 13.0 |
| <u>*First attention at NIMH*</u> |  |  |
| Before April 15th, 2020 | 270 | 76.0 |
| After April 15th, 2020 | 85 | 24.0 |
| <u>*Mode of admission to NIMH*</u> |  |  |
| Ambulatory care | 149 | 65.1 |
| Emergency department | 77 | 33.6 |
| Nonregistered | 3 | 1.3 |
| <u>*Type of attentions during follow up*</u> |  |  |
| Ambulatory face-to-face | 896 | 33.9 |
| Ambulatory telemedicine | 1,261 | 47.7 |
| Emergency department | 434 | 16.4 |
| Ambulatory, non-specified | 34 | 1.3 |
| Others, non-specified | 17 | 0.6 |
| <u>Period of ambulatory consultation</u> |  |  |
| Before April 15th, 2020 | 2,175 | 44.5 |
| After April 15th, 2020 | 2,712 | 55.1 |
| <u>Evolution of symptoms</u> |  |  |
| Unchanged | 1,814 | 61.3 |
| Improved | 665 | 22.5 |
| Worsened | 480 | 16.2 |
| <u>Loss to follow up</u> |  |  |
| No | 238 | 66.9 |
| Yes | 118 | 33.1 |

\* Mean (standard deviation)

### Survival function

The survival rate for the full follow up time was 53.9% (95%CI: 42.0-64.4%). When stratified by the period of first appointment at the NIMH, the survival function for those who started attention at NIMH after April 15^th^, 2020, was 63.0% (95%CI: 47.6-74.9%); while the survival function for those who began treatment at NIMH before April 15^th^, 2020, was 56.2% (95%CI: 43.7-67.0%). In the latter group, the survival function for a 2-year follow-up was 77.6% (95%CI: 71.9-82.3%). Detailed information can be found in Table 2 and Figure 1.

**Table 2.** Survival function for LTFU.

| Time | At risk | Events | SF | 95%CI |
| --- | --- | --- | --- | --- |
| 0 | 0 | 32 | 1 | - |
| 0.5 | 288 | 17 | 0.9 | 0.87-0.93 |
| 1 | 248 | 23 | 0.85 | 0.80-0.88 |
| 1.5 | 194 | 8 | 0.76 | 0.71-0.81 |
| <u>2</u> | <u>168</u> | <u>8</u> | <u>0.73</u> | <u>0.68-0.78</u> |
| 2.5 | 150 | 6 | 0.69 | 0.64-0.75 |
| 3 | 132 | 4 | 0.67 | 0.61-0.72 |
| 3.5 | 76 | 5 | 0.64 | 0.58-0.70 |
| 4 | 2 | 0 | 0.54 | 0.42-0.64 |
| *Start treatment at NIMH before April 15th, 2020* |  |  |  |  |
| 0 | 0 | 19 | 1 | .-. |
| 0.5 | 242 | 10 | 0.93 | 0.89-0.95 |
| 1 | 223 | 22 | 0.89 | 0.84-0.92 |
| 1.5 | 187 | 5 | 0.8 | 0.74-0.84 |
| <u>2</u> | <u>176</u> | <u>11</u> | <u>0.78</u> | <u>0.72-0.82</u> |
| 2.5 | 150 | 6 | 0.72 | 0.66-0.78 |
| 3 | 132 | 4 | 0.69 | 0.63-0.75 |
| 3.5 | 76 | 5 | 0.67 | 0.61-0.73 |
| 4 | 2 | 0 | 0.56 | 0.44-0.67 |
| *Start treatment at NIMH after April 15th, 2020* |  |  |  |  |
| 0 | 0 | 13 | 1 | .-. |
| 0.5 | 46 | 7 | 0.81 | 0.69-0.88 |
| 1 | 25 | 1 | 0.67 | 0.52-0.77 |
| 1.5 | 7 | 0 | 0.63 | 0.48-0.75 |
| <u>2</u> | <u>1</u> | <u>0</u> | <u>0.63</u> | <u>0.48-0.75</u> |

**Figure 1.**
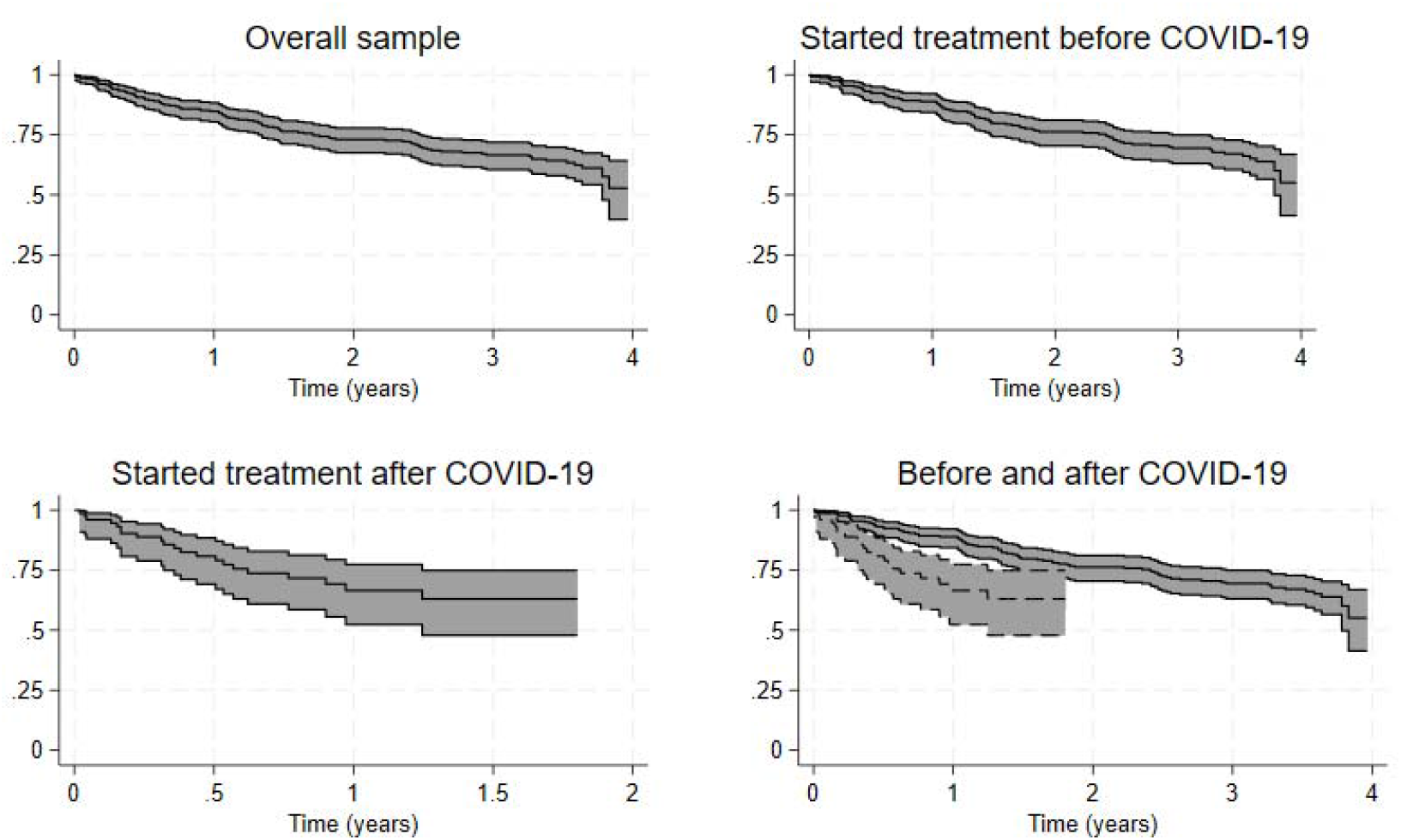
Kaplan-Meier survival estimates. The grey area corresponds to 95% confidence intervals. Central line punctual estimates.

### Bivariate analysis

Compared to those who began their attention at INSM before April 15^th^, 2020, those who did it later had 2.53 times the instantaneous risk (IR) of LTFU (95%CI: 1.55-4.51). Neither having an appointment later than April 15^th^, 2020, (HR=1.26; 95%CI: 0.80-2.04), nor the type of ambulatory attention (telemedicine vs. face-to-face) (HR=1.10; 95%CI: 0.68-1.78) were associated with LTFU. Also, compared to those treated in the psychosis program, those treated in the anxiety (HR=1.85; 95%CI: 1.03-3.33) and personality (HR=1.81; 95% CI: 0.02-3.22) programs had a statistically significant increased IR of LTFU. We found no evidence of an association between patients’ evolution from consultation to consultation or between sociodemographic variables, including sex, age, level of education, or marital status, and LTFU. Details can be found in Table 3.

**Table 3.**
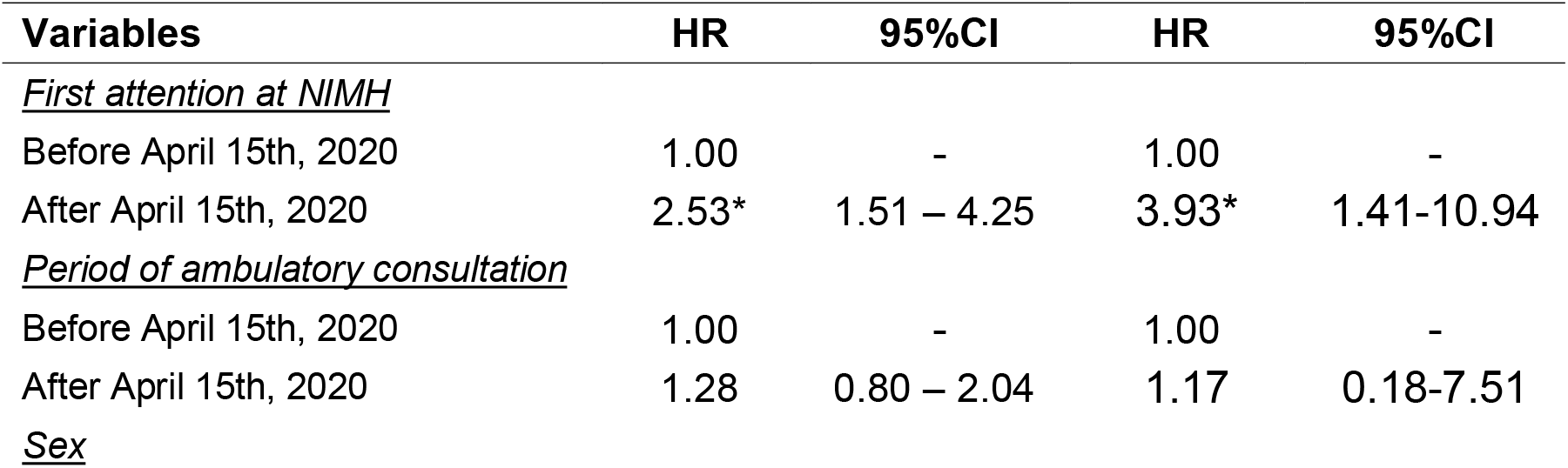
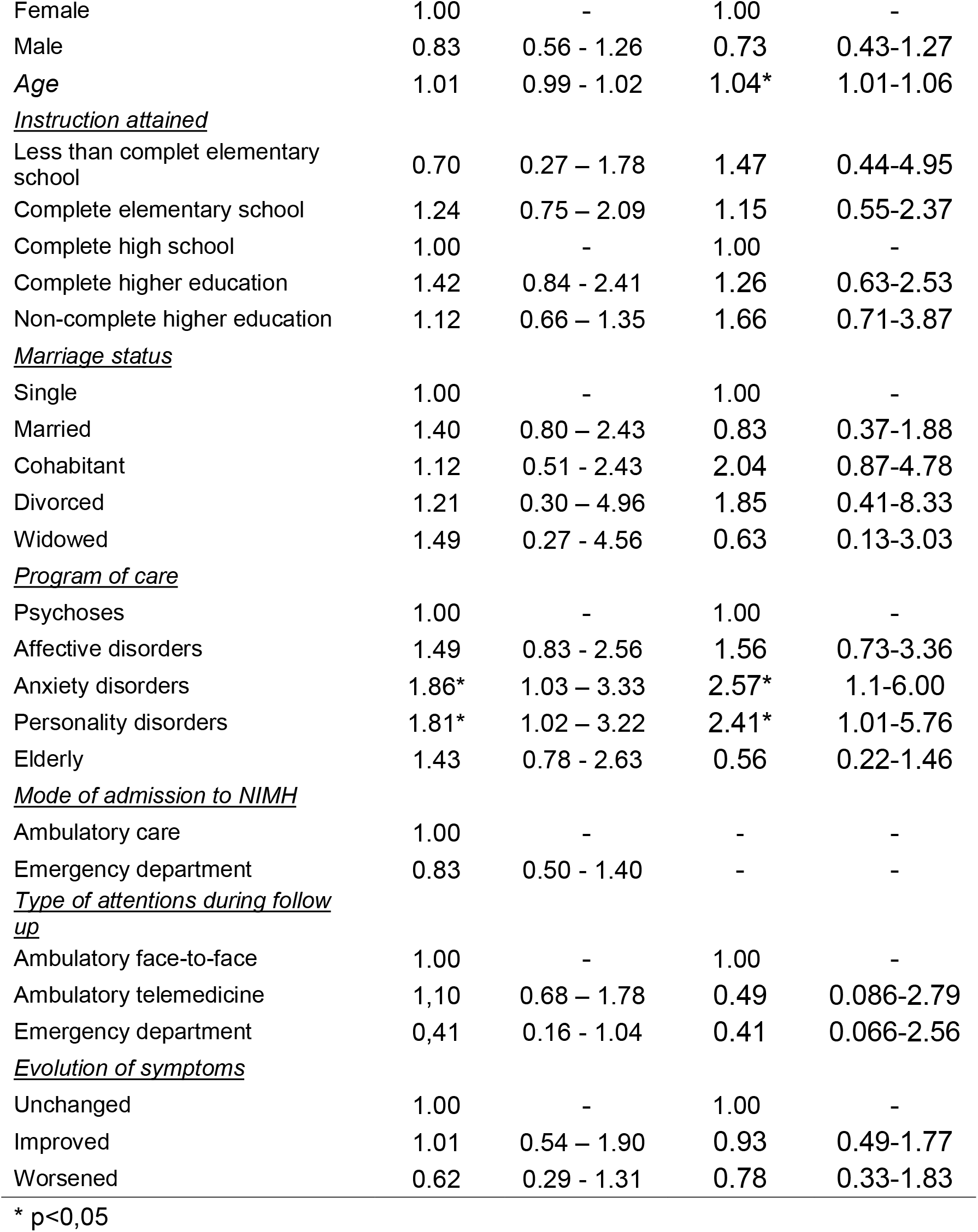
Bivariate and mulitple analysis.

**Tabla 3. Bivariate and multiple analysis**
| Variables | HR | 95%CI | HR | 95%CI |
| --- | --- | --- | --- | --- |
| *First attention at NIMH* |  |  |  |  |
| Before April 15th, 2020 | 1.00 | - | 1.00 | - |
| After April 15th, 2020 | 2.53* | 1.51 – 4.25 | 3.93* | 1.41-10.94 |
| *Period of ambulatory consultation* |  |  |  |  |
| Before April 15th, 2020 | 1.00 | - | 1.00 | - |
| After April 15th, 2020 | 1.28 | 0.80 – 2.04 | 1.17 | 0.18-7.51 |
| *Sex* |  |  |  |  |
| Female | 1.00 | - | 1.00 | - |
| Male | 0.83 | 0.56 - 1.26 | 0.73 | 0.43-1.27 |
| Age | 1.01 | 0.99 - 1.02 | 1.04* | 1.01-1.06 |
| <u>Instruction attained</u> |  |  |  |  |
| Less than complet elementary school | 0.70 | 0.27 – 1.78 | 1.47 | 0.44-4.95 |
| Complete elementary school | 1.24 | 0.75 – 2.09 | 1.15 | 0.55-2.37 |
| Complete high school | 1.00 | - | 1.00 | - |
| Complete higher education | 1.42 | 0.84 - 2.41 | 1.26 | 0.63-2.53 |
| Non-complete higher education | 1.12 | 0.66 – 1.35 | 1.66 | 0.71-3.87 |
| <u>Marriage status</u> |  |  |  |  |
| Single | 1.00 | - | 1.00 | - |
| Married | 1.40 | 0.80 – 2.43 | 0.83 | 0.37-1.88 |
| Cohabitant | 1.12 | 0.51 - 2.43 | 2.04 | 0.87-4.78 |
| Divorced | 1.21 | 0.30 – 4.96 | 1.85 | 0.41-8.33 |
| Widowed | 1.49 | 0.27 - 4.56 | 0.63 | 0.13-3.03 |
| <u>Program of care</u> |  |  |  |  |
| Psychoses | 1.00 | - | 1.00 | - |
| Affective disorders | 1.49 | 0.83 - 2.56 | 1.56 | 0.73-3.36 |
| Anxiety disorders | 1.86* | 1.03 – 3.33 | 2.57* | 1.1-6.00 |
| Personality disorders | 1.81* | 1.02 – 3.22 | 2.41* | 1.01-5.76 |
| Elderly | 1.43 | 0.78 - 2.63 | 0.56 | 0.22-1.46 |
| <u>Mode of admission to NIMH</u> |  |  |  |  |
| Ambulatory care | 1.00 | - | - | - |
| Emergency department | 0.83 | 0.50 - 1.40 | - | - |
| <u>Type of attentions during follow up</u> |  |  |  |  |
| Ambulatory face-to-face | 1.00 | - | 1.00 | - |
| Ambulatory telemedicine | 1,10 | 0.68 – 1.78 | 0.49 | 0.086-2.79 |
| Emergency department | 0,41 | 0.16 - 1.04 | 0.41 | 0.066-2.56 |
| <u>Evolution of symptoms</u> |  |  |  |  |
| Unchanged | 1.00 | - | 1.00 | - |
| Improved | 1.01 | 0.54 – 1.90 | 0.93 | 0.49-1.77 |
| Worsened | 0.62 | 0.29 - 1.31 | 0.78 | 0.33-1.83 |

\* p<0,05

### Exploratory and multiple analyses

In the multiple regression analysis, we found that, after controlling for sex, age, instruction attained, marriage status, program of care, mode of admission to NIMH, type of attention and the clinical evolution of symptoms, the magnitude of the association between LTFU and beginning attentions at NIMH after the institution of telemedicine remained statistically significant (HR=3.93; 95% CI: 1.41-10.94). Additionally, older age (HR=1.04, 95% CI: 1.01-1.06) and receiving treatment in the anxiety (HR=2.57, 95% CI: 1.10-6.00) and personality disorders programs (HR=2.41, 95% CI: 1.01-5.76) increased the IR of LTFU (See Table 3)

Using stratification, potential interaction effect between the period in which treatment at NIHM started and the type of attention (telemedicine/face-to-face/emergency) was performed. We found that among the population who started treatment before April 15^th^, 2020, both receiving telemedicine care (HR=0.42; 95% CI: 0.20-0.86) and emergency consultations (HR=0.25; 95% CI: 0.85-0.74) reduced the IR for LTFU, compared to face-to-face attention. No statistically significant differences were found between face-to-face and telemedicine or emergency care in the group that started treatment after April 15^th^, 2020 (HR=0.41; 95% CI: 0.50-3.06).

When the association between the program of care and LTFU was stratified by the period of first attention at NIMH, no statistically significant differences in IR of LTFU was found among those who began care at NIMH before April 15^th^, 2020. However, in the group that started care afterwards, a higher IR of LTFU was found in those receiving treatment for affective disorders (HR=8.91; 95% CI: 1.09-72.82) and anxiety (HR=12.16; 95% CI: 1.41-104.62) programs compared to participants in the psychosis group.

Then, we explored the association between program of care and LTFU. This association was first stratified by type of attention, where, only in the telemedicine use group a higher IR for LTFU was found in the anxiety (HR=3.03; 95% CI: 1.33-6.92) and personality (HR=2.68; 95% CI: 1.17-6.16) programs, when compared to the psychosis group. Further, when stratifying by time when attention at NIMH started, we found that in the group that started treatment at NIMH after April 15^th^, 2020, and received ambulatory care using telemedicine, those in the anxiety (HR=13.70; 95% CI: 1.59-118.23) and personality (HR=8.43; 95% CI: 1.01-70.21) programs had a significantly higher risk of LTFU.

Finally, a sub-analysis of the profile of the population by the date of first attention at NIMH, showed that those that had their first attention after April, 15^th^ 2020, were on average younger, with a higher tendency toward having completed at least high school degree of education, more likely to be treated in the affective and personality disorder programs, and have been classified as a favorable evolution. The overall proportion of LTFU was not found statistically different between these groups. Details can be found in Table 4.

**Table 4.**
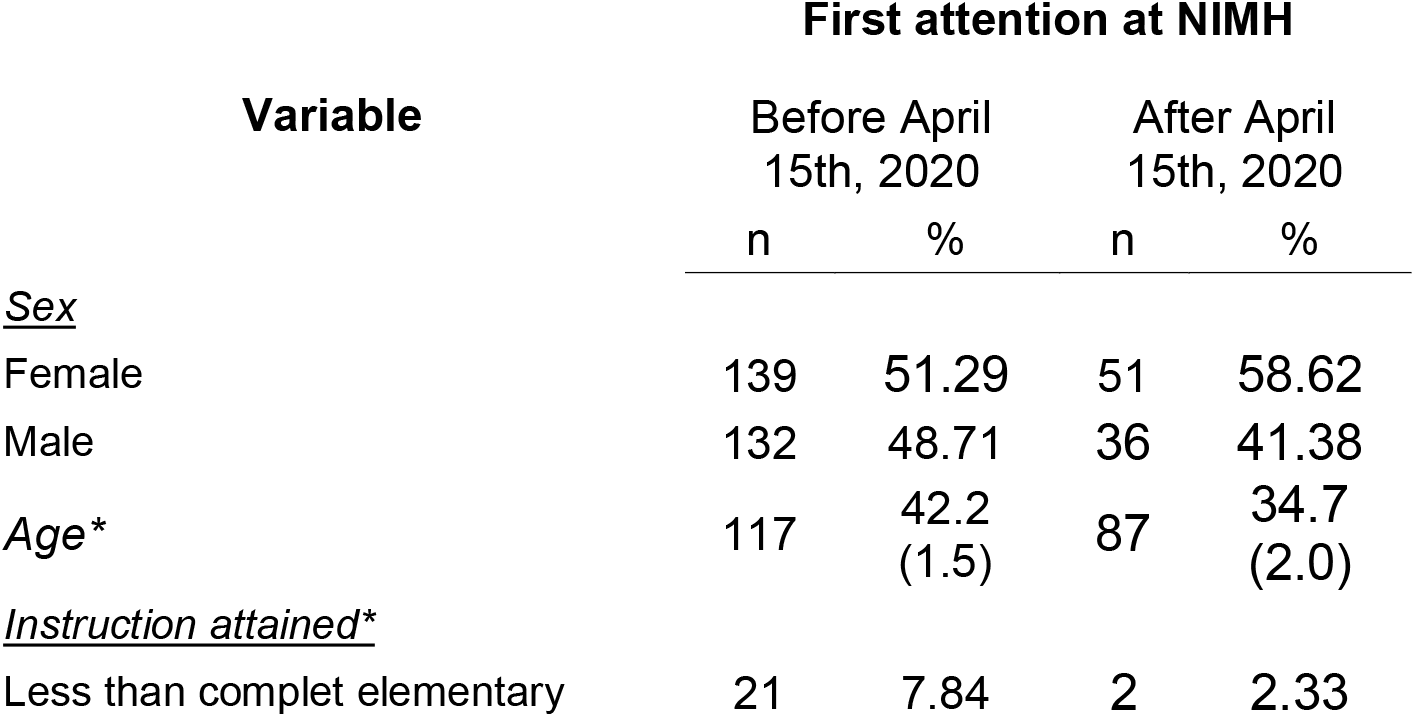
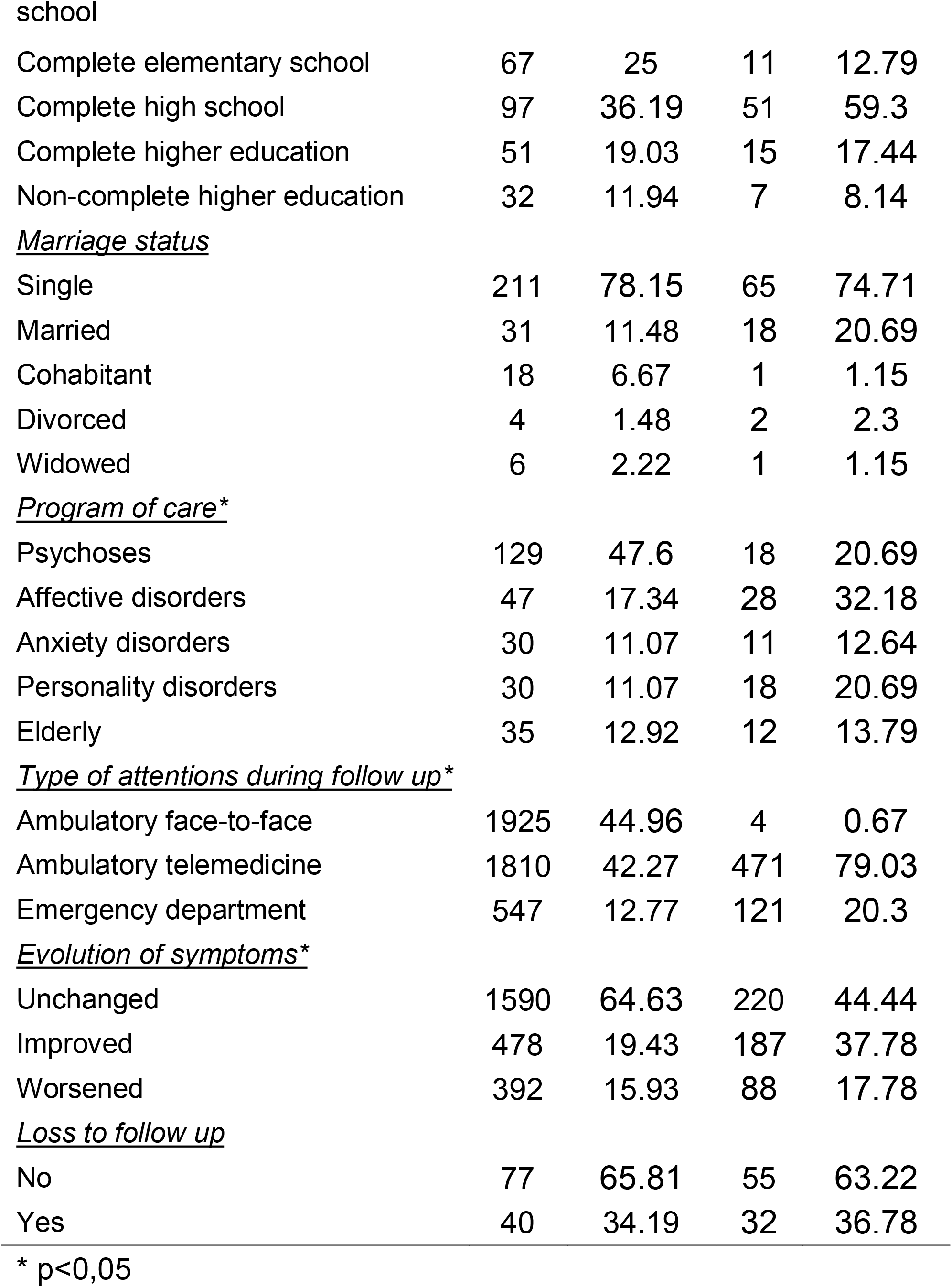
Differences in sociodemographic and clinical variables by First attention at NIMH.

## Discussion

### Key findings

Briefly, the main results of this study were that survivor function for LTFU after 2 years of follow up seems to be higher for those that began their attention at NIMH before April 15^th^, 2020, (0.78) than for those that started afterwards (0.75), while the survival function for the 4-years follow up was estimated to be 0.54. Overall, LTFU was found to be associated with beginning treatment at NIMH before April 15^th^, 2020; and to receiving care in anxiety and personality programs, compared to being at the psychosis. Further analyses suggested that in the group that started treatment at NIMH before April 15^th^, 2020, receiving telemedicine attention slightly decreased the IR of LTFU; while in the group that began treatment at NIMH afterwards, a higher IR for LTFU was found for those in the affective and anxiety disorders programs, when compared to those in the psychosis program. Finally, the subgroup of patients that started treatment at NIMH after April 15^th^, 2020, and received telemedicine attentions showed a markedly higher IR for LTFU in those in the anxiety and personality programs, when compared to those in the psychosis program. No association between the mode of attention (telemedicine/face-to-face), or the period of attention (Before or after April 15^th^, 2020) was found.

Before more detailed discussion of these findings, several limitations must be considered. Of central concern is the definition of the outcome variable. For the purpose of this study, LTFU was considered when an individual with at least one complete attention with a psychiatrist at NIMH during the follow up period, did not have a follow-up psychiatric appointment afterwards; however, it is not possible to determine whether this lack of clinical follow up was due to clinical improvement, a change of healthcare provider without formal referral, or the patient’s death (when a referral or information of death was recorded in the clinical chart, it was not considered LTFU). Also, our sample size might not allow to detect mild to low effect size associations, particularly in the variables with multiple categories.

The analyzed data do not suggest that having the attention delivered by telemedicine in contrast to face-to-face, nor having an ambulatory appointment before or after April 15^th^, 2020, increases the risk for LTFU in the overall sample. However, starting to receive mental healthcare at the NIMH before or after April 15^th^, 2020, might be an important variable to understand the behavior of LTFU. On this date and forward, all psychiatric ambulatory attentions for adults at the NIMH started being delivered exclusively via telemedicine, due to a series of measures designed to adapt to the COVID-19 pandemic in Peru. During this period a substantial part of lower-level mental healthcare centers in Lima suffered a decrease in the number of attentions starting from March, where the first case of COVID-19 was detected in Lima, recovering roughly 9 months later, by November 2020. At the same time, a possible increase of depressive and anxiety symptoms was described in Peru, mirroring the reporting globally, with an excess of demand for mental healthcare [4–8].

These epidemiological phenomena, including the reduction in the offer of specialized healthcare in the lower levels of the health system, coinciding with an increment of demand might have shifted the profile of persons starting attention at NIHM, and thus modifies the risk for LTFU. For example, our results suggest that the proportion of persons with affective disorders increases from 18.5% in the period before April 15^th^, 2020, to 28.9% afterwards; with a similar in magnitude decrease in the proportion of persons with psychosis. Furthermore, the IR for LTFU might behave differently in both groups, with evidence of a lower IR in persons receiving attention by telemedicine in comparison to face-to-face, for those in the group that began treatment at NIMH before April 15^th^, 2020, and an increased risk for those beginning afterwards.

These differences might be explained by divergence in the severity and evolution of symptoms of individuals that started being treated at NIMH after April 15^th^, 2020, thus, due to the saturation of healthcare centers with capacity to treat mental disorders due to the COVID-19 pandemic, patients with milder forms of affective disorders, mainly depressive and anxiety disorders, might have starting seeking attention at NIMH, and recovered as social, work and health conditions improved. Also, as the capacity for attention of the lower-level mental health centers returned to pre-pandemic levels, as *Villareal-Zegarra et al*. described, it is feasible that patients stopped their appointments at NIMH without any communication; this would be perfectly possible as formal reference needs to be made from lower-level mental health centers to the NIMH, but not vice-versa, and no formal obligation of communicating the continuation of treatment exists for the patients or the lower-level mental healthcare centers [24].

### Implications

Our results show that the use of telemedicine as a delivery tool for mental health care is not associated with a higher risk of LTFU, except in the group of patients that started their treatment at NIMH during the COVID-19 pandemic. This latter group has been shown to present distinct clinical characteristics, with a higher proportion of affective disorders, that might not be consistent with the usual profile of individuals treated at a specialized institution. In future research that builds from these findings, it should be necessary to study LTFU in a post COVID-19 scenario and to identify sub-populations of patients that benefit the most from the use of telemedicine and face-to-face attention; and the feasibility of procedures directed toward early identification and profiling of LTFU.

## Data Availability

All data produced in the present study are available upon reasonable request to the authors

